# EVALUATION OF THE ABBOTT SARS-COV-2 IG-G ASSAY

**DOI:** 10.1101/2020.06.28.20132498

**Authors:** CS Lau, SP Hoo, YL Liang, TC Aw

## Abstract

**Introduction:** Antibodies to the novel severe acute respiratory syndrome coronavirus 2 (SARS-CoV-2) can increase as soon as 10-13 days after infection. We describe our evaluation of the Abbott SARS-CoV-2 IgG assay on the Architect immunoassay analyser.

**Methods:** We assessed the precision, sensitivity, and specificity of the Abbott SARS-CoV-2 IgG assay in samples from polymerase chain reaction (PCR) positive patients and healthy healthcare workers. The manufacturer cut-off index (COI) of 1.4 was adopted to identify positive results. We examined the assay cross-reactivity with other viral antibodies (influenza/dengue/hepatitis C/hepatitis B) and rheumatoid factor (RF). The sample throughput of the Abbott assay was also assessed.

**Results:** The Abbott assay showed excellent precision, with a CV of 3.4% for the negative control (COI = 0.06) and 1.6% for a high positive serum sample (COI = 8.6). Residual serum was available from 57 inpatients not initially suspected of having COVID-19, 29 of whom tested positive for SARS-CoV-2 IgG. The Abbott assay has a sensitivity of 90.9-100% when tested in 54 subjects ≥14 days post PCR positive, and a specificity of 100% (N = 358). There was no cross-reactivity with other viral antibodies (influenza/dengue/hepatitis C/hepatitis B) and RF. The Architect Abbott assay has a throughput of 100 samples in 70 minutes.

**Conclusion:** The Abbott SARS-CoV-2 IgG assay shows excellent performance that is well within FDA and CDC guidelines when testing patients ≥14 days POS with little cross-reactivity from other viral antibodies. There is some evidence that SARS-CoV-2 IgG develops early in the disease process.

**IMPACT STATEMENT:** With the current SARS-CoV-2 pandemic still ongoing, laboratories are hard pressed to introduce SARS-CoV-2 antibody testing to help as an indirect marker for infection to identify patients with prior infection/exposure. The Abbott SARS-CoV-2 IgG assay has excellent performance with good precision, specificity, and sensitivity ≥14 days after a positive SARS-CoV-2 PCR test, with a competitive throughput of 100 samples in 70 minutes. It shows no cross-reactivity with other viral antibodies (influenza/dengue/hepatitis B/hepatitis C) and rheumatoid factor. We have also found evidence of early antibody development in some patients before they tested positive on the SARS-CoV-2 PCR test.

## INTRODUCTION

The novel severe acute respiratory syndrome coronavirus 2 (SARS-CoV-2) can lead to mild or moderate coronavirus disease 2019 (COVID-19) with cough, fever, malaise, myalgias, gastrointestinal symptoms and anosmia (1), or a severe disease with acute respiratory distress syndrome (2). The need for the serological diagnosis of SARS-CoV-2 has become acute. The Centers for Disease Control and Prevention (CDC) (3) and the Royal College of Pathologists of Australasia (RCPA) (4) stipulate that only viral testing (nucleic acid or antigen) is the standard test to diagnose acute infection. However, some studies (5, 6) show that the SARS-CoV-2 antibodies can increase as soon as 10-13 days after infection. Antibody tests can act as an indirect marker for infection and help identify patients with prior infection/exposure who have developed an immune response (7). Abbott has developed a SARS-CoV-2 IgG assay for use with its Architect immunoassay analysers for the rapid testing of large numbers of samples. We describe our evaluation of the Abbott SARS-CoV-2 IgG assay.

## MATERIALS AND METHODS

### Patient cohorts

Residual serum from cases with suspected or confirmed SARS-CoV-2 infection from April to May 2020 were recruited. Samples testing positive for dengue fever/Anti-HCV/HBsAg/anti-HBc IgM and rheumatoid factor (RF) were used to check for cross-reactivity in the SARS-CoV-2 IgG assay. Volunteers (laboratory staff and frontline healthcare workers) from our hospital also provided serum samples as control subjects. All samples were collected in plain serum tubes.

### Instrumentation and analysis

For SARS-CoV-2 polymerase chain reaction (PCR) testing, our hospital molecular laboratory employs a duplex real-time PCR that targets the N and E genes using a Qiagen EZ1 extraction system and Rotor Gene Q amplification system. The Abbott SARS-CoV-2 IgG assay is a qualitative chemiluminescent microparticle immunoassay used for the detection of IgG antibodies to SARS-CoV-2 (undisclosed epitope on the viral nucleocapsid) in human serum/plasma on the ARCHITECT i2000 System. The sample, SARS-CoV-2 antigen coated paramagnetic microparticles, and assay diluent are combined and incubated. Thereafter, anti-human IgG acridinium-labelled conjugate is added to create a reaction mixture. Following the addition of pre-trigger and trigger solutions, the resulting chemiluminescent reaction is directly proportional to the amount of IgG antibodies. When compared to the mean chemiluminescent signal of a calibrator, an IgG index is derived with a reported cut-off index (COI) of 1.4. The assay has a stated inter-assay precision (CV) of 5.9% and 1.2% for negative and positive controls at a mean COI of 0.04 and 3.53. The reported assay specificity is 99.6% and sensitivity of 100% for samples ≥14 days post symptom onset. To prevent potential interactions, maintenance has to be performed to and following the batching of 50 SARS-CoV-2 IgG samples (Abbott SARS-CoV-2 IgG assay package insert, H07891R02, April 2020).

Statistical analyses were performed using MedCalc software v19.3.1 (MedCalc, Ostend, Belgium). As this work was part of routine evaluation of new diagnostic assays, it was deemed exempt by our institutional review board.

## RESULTS

For precision analysis, 5 pools (including negative and positive Abbott controls, and 3 positive serum samples over a range of COI values) were run 5 times daily over 5 days as per the CLSI EP15-A3 protocol (8). The Abbott assay showed excellent precision, with a CV of 3.4% (negative control, COI = 0.06) and 1.6% (serum sample, COI = 8.6) (Supplemental Table 1).

For sensitivity testing [positive percentage agreement, (PPA)], we used 338 PCR positive COVID-19 cases; cases with unknown PCR status (N = 5) and PCR negative cases (N = 10) were excluded. 57 of these cases were residual samples from inpatients who were not initially suspected of having COVID-19 but subsequently tested positive for SARS-CoV-2 PCR. Notably, 29 of these 57 samples were positive for IgG prior to having a positive PCR test. Of the remaining 266 samples, the PPA increased to 90.9% after 14 days post positive PCR (POS) and 100% by 21 days POS (see Table 1).

**Table 1:** Abbott assay sensitivity (positive percentage agreement) by days post positive PCR.

| Days POS | N | IgG positive | IgG negative | PPA (%) | 95% CI |
| --- | --- | --- | --- | --- | --- |
| 0-<7 | 155 | 76 | 79 | 49.03 | 40.93 to 57.18 |
| 7-<14 | 57 | 43 | 14 | 75.44 | 62.24 to 85.87 |
| 14-<21 | 22 | 20 | 2 | 90.91 | 70.84 to 98.88 |
| ≥21 | 32 | 32 | 0 | 100.00 | 89.11 to 100.00 |

For specificity testing [negative percentage agreement (NPA)], we included 294 samples from healthcare workers, 46 samples positive for dengue, 3 samples positive for Anti-HCV, 8 samples positive for HBsAg, 2 samples positive for anti-HBc IgM, and 5 RF positive samples. All these samples were from subjects with no suspicion for COVID-19. None of these cases tested positive, yielding a total specificity of 100% (95% CI 98.98 to 100.00) (See Table 2). The sero-negativity of our healthcare workers also underscores the adequacy of our personal protective equipment measures. However, they remain at risk for future COVID-19 infections, and continued safety precautions should still be enforced. We also analysed for any potential cross-reactivity from antibodies consequent to recent influenza vaccination (239 staff had received their influenza vaccination 4 weeks prior to testing). None of these cases (dengue, recent influenza vaccination cases, or positive Anti-HCV, HBsAg, anti-HBc IgM and RF) tested positive for SARS-CoV-2 IgG.

**Table 2:** Abbott assay interference analysis and specificity (negative percentage agreement).

|  | N | Abbott negative | Abbott positive |
| --- | --- | --- | --- |
| Healthcare workers | 294 | 294 | 0 |
| Dengue fever | 46 | 46 | 0 |
| Anti-HCV | 3 | 3 | 0 |
| HBsAg | 8 | 8 | 0 |
| Anti-HBc IgM | 2 | 2 | 0 |
| RF positive | 5 | 5 | 0 |
| Total | 358 | 358 | 0 |
| Abbott NPA | 100% (95% CI 98.98 to 100.00) |  |  |

The throughput of the Architect Abbott assay was examined. The Architect was able to analyse 100 specimens for SARS-CoV-2 IgG in 1 hour 9 minutes and took 2 hours 9 minutes to analyse 250 specimens.

## DISCUSSION

The Abbott SARS-CoV-2 IgG assay shows excellent performance, with a CV of 3.4% and 1.9% for the negative and positive calibrators. The diagnostic sensitivity of this assay is 90.9-100% ≥14 days POS, with a specificity of 100%. This is in close agreement with the manufacturer’s specifications. Our results are also in keeping with other studies (9-12) that have evaluated the Abbott SARS-CoV-2 IgG assay, where the sensitivity/PPA increases to >90% after 14 days post symptom onset. The US CDC and the Food and Drug Administration (FDA) have stipulated a sensitivity of 90% and specificity of 95% for an acceptable SARS-CoV-2 serology test (13, 14). The Abbott assay easily falls within these requirements. Furthermore, the Abbott assay has an acceptable throughput, which is comparable to other SARS-CoV-2 immunoassays. In the latest revision of the package insert (Abbott SARS-CoV-2 IgG package insert, H70891R03, May 2020), it is heartening to note that the requirement for batch analysis and maintenance pre- and post-testing has been removed.

In our study we have used days POS to categorize our population. Using the onset of patient symptoms as a criterion for SARS-CoV-2 classification will result in the exclusion of asymptomatic patients (15, 16). Using days POS mitigates against this pitfall. We also found 29 patient samples that tested positive for SARS-CoV-2 IgG before a positive PCR test. This underscores the fact that SARS-CoV-2 antibody development may occur earlier than expected in some cases. Our study also confirms that the assay remains specific even in patients with previous dengue infection, recent influenza vaccination, or in the presence of Anti-HCV, HBsAg, anti-HBc IgM and RF. A recent report (17) found that SARS-CoV-2 infection may cause false-positive dengue antibody results. It is noteworthy that positive dengue serology in 46 patients did not cause a false positive SARS-CoV-2 IgG. Other evaluations of the Abbott assay also demonstrate little cross-reactivity in patients with previous viral infections and haemodialysis (9). Finally, we have also provided data on the sample throughput of the Abbott assay. A limitation of our study is that it is a single centre study, and we do not have the sero-prevalence of SARS-CoV-2 in our community population. In addition, we only had few samples for cases before and after 0-<7 days POS. Further studies on larger populations would be desirable.

## Conclusion

Overall, our results show that the Abbott SARS-CoV-2 IgG assay has excellent performance and is highly comparable both with the manufacturer’s information and other published studies. We look forward to larger studies over longer periods of time to further evaluate the use of serological assays in the management of COVID-19.

## Data Availability

nil

## ABBREVIATIONS

SARS-CoV-2: Novel severe acute respiratory syndrome coronavirus 2
COVID-19: Coronavirus disease 2019
CDC: Centers for Disease Control and Prevention
RCPA: Royal College of Pathologists of Australasia
RF: Rheumatoid factor
PCR: Polymerase chain reaction
COI: Cut-off index
CV: Inter-assay precision
CAP: College of American Pathologists
PPA: Positive percentage agreement
POS: Post positive PCR
NPA: Negative percentage agreement
FDA: Food and Drug Administration

## Notes

### Competing Interest Statement

The authors have declared no competing interest.

### Author Declarations

Changi General Hospital Institutional Review Board

